# Impact of training modules on physicians’ perspective of COVID-19: An online survey

**DOI:** 10.1101/2021.01.13.21249689

**Authors:** Badar Uddin Umar, Nazmun Nahar Alam, Tanbira Alam, Mahmudul Mannan, S M Niazur Rahman

## Abstract

**Background:** The outbreak of COVID-19 has remained a massive challenge for healthcare workers specially physicians. Effective professional training has a crucial role in preparing doctors for responding to pandemics.

**Objective:** To assess the effectiveness of existing training modules on enhancing knowledge, ensuring safe practice, and improving behavior on COVID-19 among physicians.

**Methods:** This is a descriptive, cross-sectional, online survey; where a virtual questionnaire was used to collect data through online professional platforms. A pre-tested survey tool was employed to assess the impact of professional training on infection prevention and control.

**Results:** Total 161 physicians participated in this survey from 15 different countries. Most of the respondents (72%) received training from various sources like the workplace (60%) and international agencies (21%), through the in-person or online format. Knowledge assessment revealed advanced (43%) and competent (40%) understanding by the participants. Improving knowledge progression was displayed by the cohort who received professional training (p<0.00). Physicians’ positive behavior and good practices were observed with the training modules.

**Conclusion:** It became evident from this study, that professional training is effective in enhancing knowledge, improving behavior, and ensuring safe practices. Hence, designing such training modules for the physicians is warranted to tackle ongoing and future pandemics.

## 1. Introduction

The novel coronavirus (COVID-19) is a global threat since it was identified in late 2019^1^. COVID-19 infection is a highly contagious disease and has affected a large population, the total number of deaths caused by this virus has exceeded any of its predecessors. As of early November 2020, more than 49 million confirmed cases, and one million confirmed deaths across the globe, as reported by the WHO^2^. The first wave of the outbreak took place between March and July 2020. The world is clearly in the grip of a second wave of COVID-19 since July-August, with no imminent vaccine in sight^3, 4^. This pandemic has instigated a global economic downturn regardless of the income level of the country^5^. It is essential to provide the safety of healthcare workers not only to sustain continuous patient care but also to ensure they do not transmit the virus^6^. The medical community worldwide was confronted with the need to treat a new, unknown disease, where patients’ management was challenging as current guidelines are lacking and some of the used therapies are only addressed to patients included in trials^7, 8^. Medical information is rapidly and continuously being updated as new data are available while fighting this emerging disease^8^.

The COVID-19 outbreak is a huge challenge for physicians, who need strong comprehensive skills to deal with it effectively. Continuing medical education has an imperative role in preparing for and responding to such emergencies. To mitigate the outbreak and defy the pandemic, different online courses for healthcare workers around the world were initiated; funds were raised globally; strategic preparedness and response plan (SPRP) was set up aimed to protect the health care systems^7^. In this era of physical distancing, numerous online training for physicians provides an additional advantage of instant knowledge acquisition, flexible learning, and rapid management of patients. Increasing the level of knowledge and proper training of physicians can significantly curb the development and transmission of disease to other patients and health care workers (HCW). Therefore, hospitals that provide services to patients with suspected or confirmed COVID-19 should aim to increase the level of knowledge of HCW and provide them with high-level training^9^.

Though there are many training modalities available in various formats, we have scanty reports on their effectiveness. The objective of this study was to assess the impact of existing training modules on enhancing knowledge, ensuring safe practice, and improving behavior among the physicians. Our findings would increase awareness to gain professional knowledge through Infection Prevention and Control (IPAC) training and prepare physicians for COVID-19’s next waves to come.

## 2. Methods

An observational, cross-sectional, web-based study was conducted among physicians who are licensed to practice in different countries by using a validated structured questionnaire in September 2020; when the world has experienced the first wave of COVID-19 infection and waiting for proper vaccination.

A questionnaire was developed containing 31 items by using WHO guidance document^10^, previously used survey tools^11, 12^, and web-based materials^13^ on emerging respiratory viruses, including COVID-19. The survey covered physicians’ characteristics, awareness, information sources, knowledge, behavior, and practices related to COVID-19. The draft survey instruments were created using Google^©^ form and were made accessible through a web link to 15 experts from different geographic regions to assess its consistency, clarity, relevance, and acceptability. Another seven (7) physicians evaluated the feasibility of the survey as well as assessed the approximate time to finish. Those responses were not included in the final analysis of the research project.

Refinements were made as needed to encourage a clearer understanding and organized the questions before the final survey was circulated via a Web link to the targeted population. The 31-item survey took approximately 15 minutes to complete. The survey instruments were divided into four parts having questions on participants’ information (13 items), and knowledge (08 items), behavior (05 items), practice (05 items) related to COVID-19.

The information section included questions on general demographic, knowledge was evaluated by multiple-choice questions focusing on etiology, signs and symptoms, transmission, and risk avoidance of COVID-19. Each response was scored either as correct or wrong, with total scores ranging from 0 to 8. A cut-off level of <4 was considered to indicate poor knowledge about COVID-19, whereas 4-6 was deemed to be competent and >6 as advanced. Behavior and practices toward COVID-19 were assessed using 05 items in each section, and each question was labeled as “Yes” or “No”; scores ranged from 0 to 5.

### Data Analysis

The collected data were validated, coded, and analyzed using SPSS version 24 (IBM). Descriptive analysis was applied to calculate frequencies and proportions. The chi-square test was used to investigate the level of association among variables. A p-value of less than 0.05 was considered statistically significant.

### Ethical Consideration

The institutional review and approval for the study protocol were obtained (Ref: HFRCMC-IRC/2020.09.050). Throughout the analysis, the confidentiality of personal information was protected by keeping the data of participants anonymous and asking participants to provide truthful answers. The involvement of qualified physicians in this survey was voluntary and was not paid. Electronic informed consent was shown on the initial page of the survey. The study was performed following the Declaration of Helsinki, as revised in 2013.

### Study Subjects

A convenience purposive sampling technique was used to select participants. The survey link was circulated to the physicians through verified online professional and social media platforms. A total of one hundred sixty-one (161) physicians participated in this study from different countries worldwide and responded voluntarily. During the data collection process, participants confidentially were strictly and adequately maintained, and all collected data were double-checked, sorted, and cleaned for consistency.

## 3. Results

### Overview

A total of 161 physicians participated, including 128 (79%) men and most of them were either in the age group of 25-35 years (35%) or 46-55 years (37%). Doctors from 15 different countries responded to our survey and the majority belonged to lower-middle-income countries 114 (71%). Professionally, most of our participants were in early career stage 69 (43%), working primarily in outpatient setup (39%). More than one-third of the respondents were directly involved in dealing with known COVID-19 cases, and about half of them (n=25) were diagnosed positive for novel coronavirus infection. Table 1 shows the sociodemographic details and table 2 summarized the professional and COVID-19 related information of the participants.

**Table 1:** Socio-demographic features of respondents; n=161

| Features | Respondent |
| --- | --- |
| Gender | 128 Male, 39 Female |
| Age | 25 – 35 years: 35% |
|  | 36 – 45 years: 19% |
|  | 46 – 55 years: 37% |
|  | 56 – 65 years: 09% |
| Region of practice | High income countries: 13% |
|  | Middle income countries: 16% |
|  | Lower middle-income countries: 71% |

**Table 2:** Respondents’ professional involvement and COVID-19 related information

| Features | Respondent |
| --- | --- |
| Practice standing | 138 Yes, 23 No |
| Clinical Setting | Outpatient: 39% |
|  | Inpatient: 33% |
|  | ICU: 04% |
|  | Others: 24% |
| Direct contact with COVID-19 patients | 58 Yes, 103 No |
| Diagnosed with COVID-19 | Yes (16%) |

### Professional training information

Most of our respondents (72%) had received IPAC training from various sources, out of which the highest being from their workplace (60%) and least from international agencies (21%). The mode of training was online (32%), physical (22%), and a combination of both (47%). According to the duration, the training our participants had received was transient (38%), intermediate (30%), and intense (32%). Detailed information on training has been summarized in Table 3.

**Table 3:**
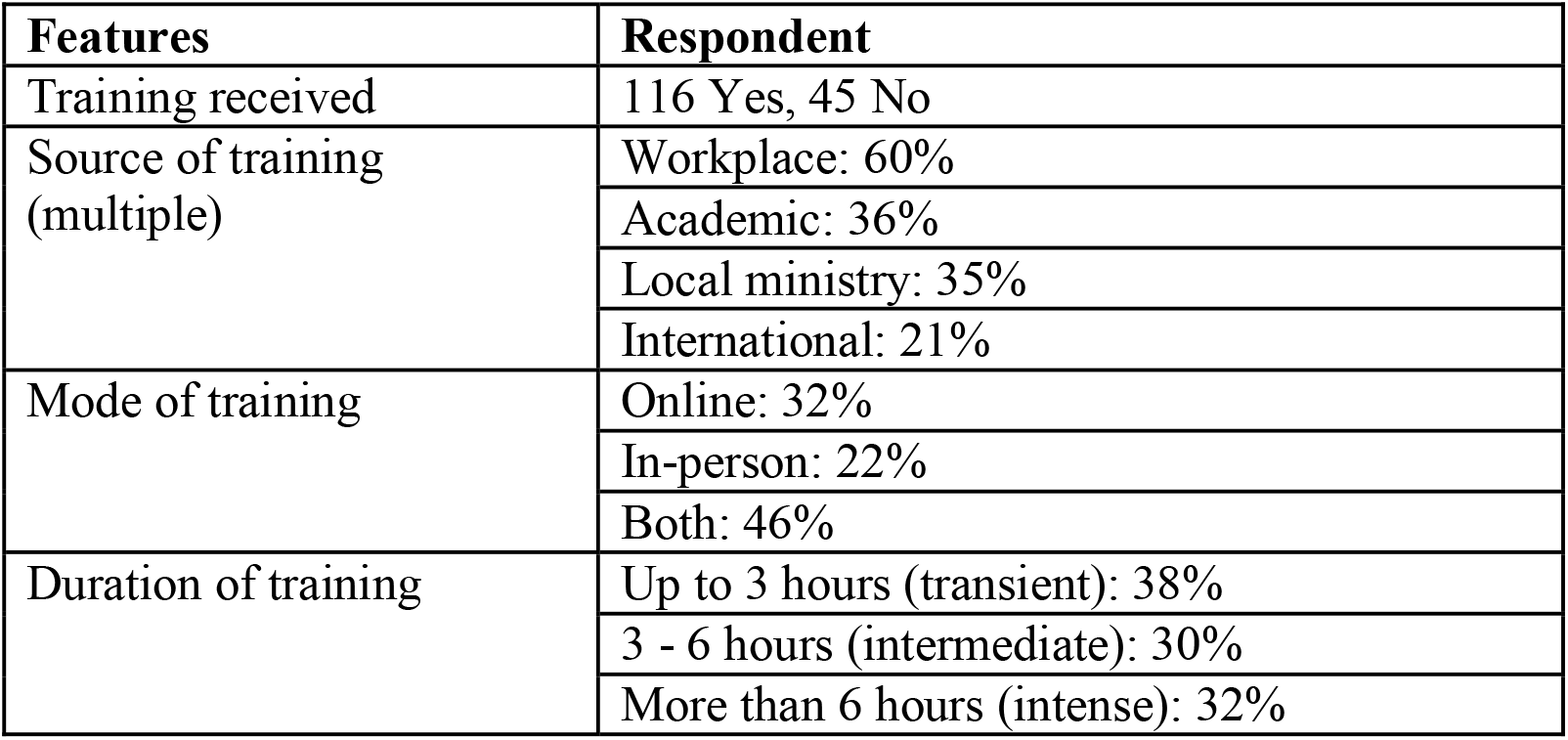
Professional training received by the participants

### Knowledge on COVID-19

Most of the participants’ overall knowledge about COVID-19 was in advanced (43%) or competent (40%) level, whereas only 16% displayed poor understanding. The pattern of correct response was sloped down with an increasing level of difficulty, i.e. less correct answers for more difficult items. Responses to the individual knowledge questions are detailed in Table 4. No knowledge differences were observed between professional levels or regions of practice.

**Table 4:**
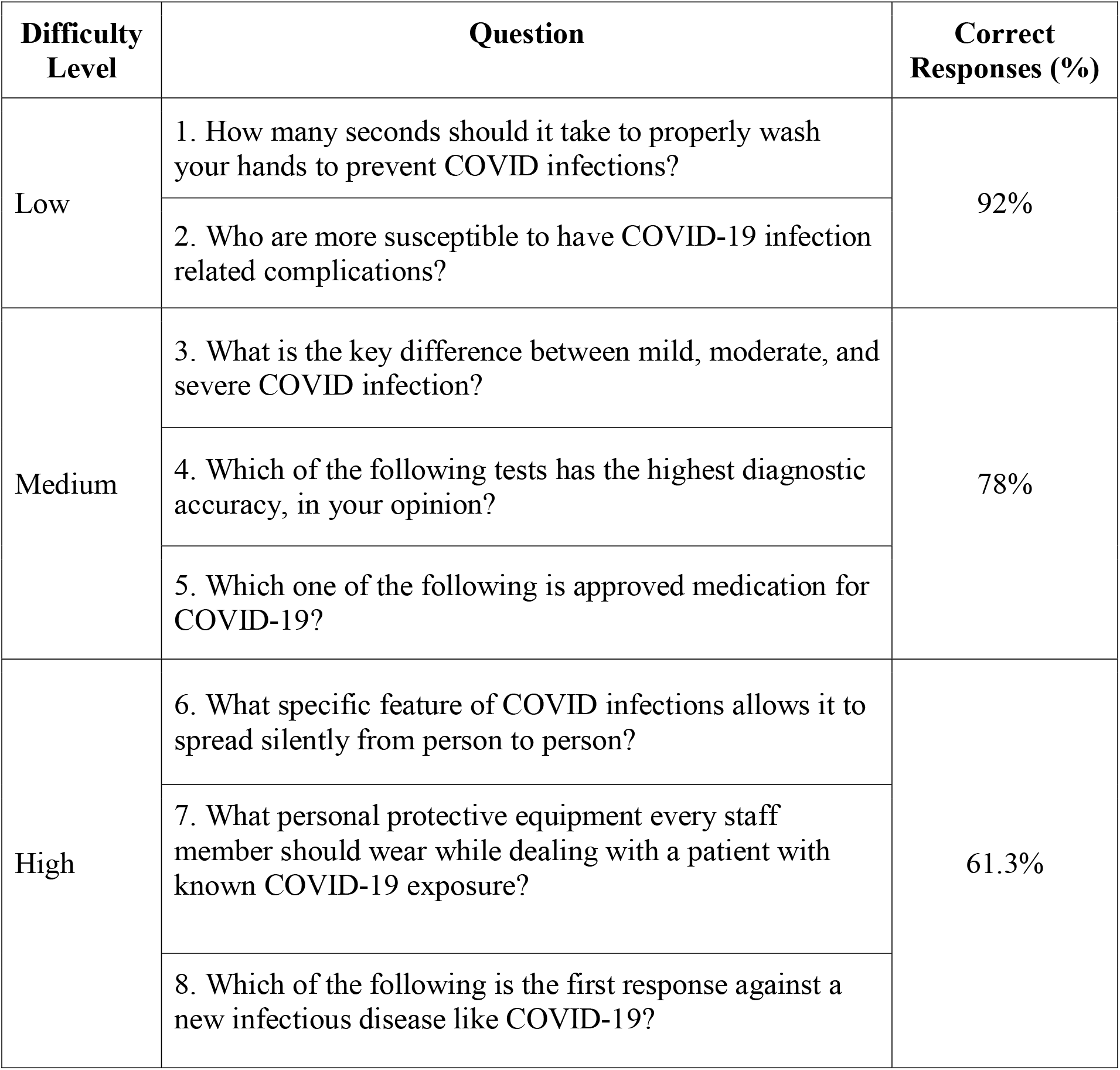
Response summary of the knowledge questions; n=161

| **Difficulty Level** | **Question** | **Correct Responses (%)** |
| --- | --- | --- |
| Low | 1. How many seconds should it take to properly wash your hands to prevent COVID infections? | 92% |
|  | 2. Who are more susceptible to have COVID-19 infection related complications? |  |
| Medium | 3. What is the key difference between mild, moderate, and severe COVID infection? | 78% |
|  | 4. Which of the following tests has the highest diagnostic accuracy, in your opinion? |  |
|  | 5. Which one of the following is approved medication for COVID-19? |  |
| High | 6. What specific feature of COVID infections allows it to spread silently from person to person? | 61.3% |
|  | 7. What personal protective equipment every staff member should wear while dealing with a patient with known COVID-19 exposure? |  |
|  | 8. Which of the following is the first response against a new infectious disease like COVID-19? |  |

### Association of professional training with knowledge on COVID-19

Participants’ IPAC training was found to be effective in the areas of knowledge progression, behavior, and practice towards COVID-19. There was an uptrend in the total knowledge among the respondents who received training, while the opposite was displayed by the non trained cohort (Figure 1). A significant relationship (p<0.000) was observed in responding to high difficulty questions by the advanced trained participants. Duration or source of training showed no differences yet, training duration had a positive impact on knowledge progression.

**Figure 1:**
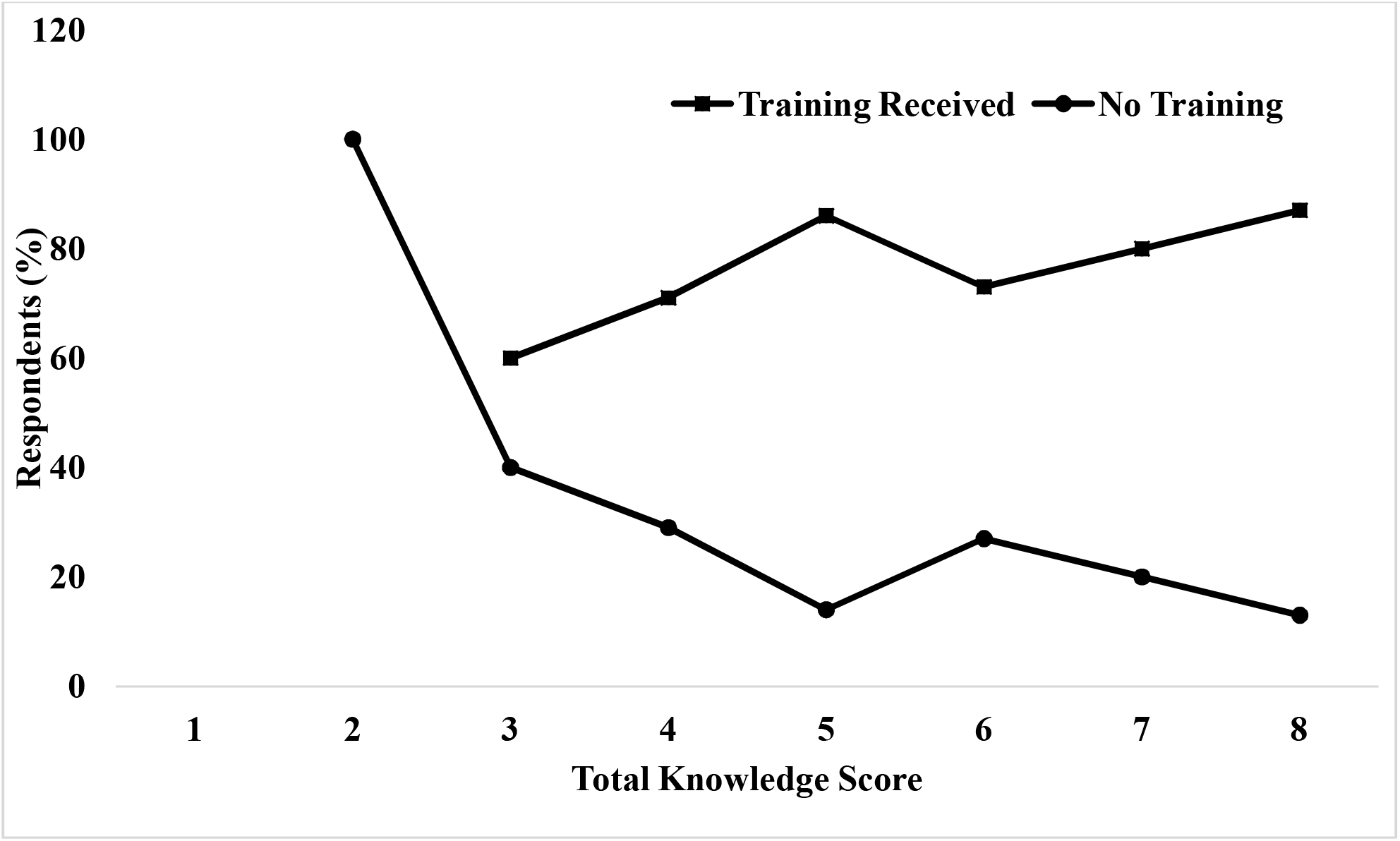
Trend of total knowledge score in relation to professional training (n=161). An upward trend of knowledge is being shown by the respondents who received the training, however, the decline of total knowledge score is displayed by the no-training recipients.

### Behavior towards COVID-19

More than 68% of the physicians exhibited a positive behavior towards COVID-19. Though statistically not significant (p=0.06), our respondents showed more positive behavior patterns in the professionally trained group. Almost similar behavioral status was observed across the clusters of participants.

### Practice about COVID-19

While dealing with COVID-19 issues, the majority of the doctors (60%) demonstrated good practice. Training has positively impacted the physicians’ practice to the disease, not significantly (p=0.06), but some extent. Almost similar practices were observed across the clusters of participants. Overall knowledge, behavior, and practice level of the respondents are displayed in Table 5.

**Table 5:** Overall knowledge, behavior, and practice of the respondents: n=161

| Features | Respondent |
| --- | --- |
| Knowledge Progression | Poor: 17% |
|  | Competent: 40% |
|  | Advanced: 43% |
| Behavioral Pattern | Positive/Good: 69% |
|  | Negative/Poor: 31% |
| Practice Trend | Positive/Good: 60% |
|  | Negative/Poor: 40% |

## 4. Discussion

The ongoing COVID-19 pandemic has put an overwhelming burden on the health systems of all the countries. To face the challenges immediate, effective, and appropriate responses along with interventions and policy changes were needed^10^. Health and safety of first-line (healthcare) workers are of great concern, as they are one of the most vulnerable groups to get exposed to infections^11^ while caring for patients. Proper IPAC training can significantly prevent the spread of infection among healthcare workers by enhancing knowledge, altering attitudes, and affecting the practice^14^.

The present cross-sectional, online survey was conducted to explore the effects of IPAC training interventions on the knowledge, behavior, and practice of physicians in a global context. This is one of the earliest among such studies conducted on physicians’ perspectives about the COVID-19 pandemic. This study was conducted toward the later phase of the pandemic (September 2020). The findings would help us to explore the causal relationship between the knowledge domain of IPAC, behavior, and practice among physicians. As well it would reveal the role of IPAC training on physician’s practices, and the risk of getting infected. Previous studies proved the efficacy of training in altering the knowledge, perception, confidence, psychosocial attitudes, and beliefs of the physicians in other clinical areas^15, 16^. Some recent studies also have suggested more focused training towards improving understanding of risks and prevention of COVID-19^12, 17^. An assessment of HCW knowledge, attitudes, and practices could provide an insight into the prevention of COVID-19 and stopping further spread of the epidemic^11, 12, 14^.

More than half (54%) of the respondents in the present survey belonged to a relatively younger age group (<25 years to 45 years). This finding is supported by previous works where the majority of respondents belonged to similar groups^18-20^. We used an online platform for reaching out to our target population, as this media is more popular among this age group of people, it was reflected through this finding.

Considering the geographical location, we had physicians from fifteen countries: Canada, USA, Brazil, Malaysia, India, Bangladesh, Pakistan, UK, Nepal, Sri Lanka, Oman, Saudi Arabia, Australia, Turkey, and UAE. We have categorized the country distribution based on the world bank’s economic data^21^ into; high-income, middle-income, and lower-middle-income groups. Though the overall health system performance index is higher for wealthy nations^22^, our study results did not show any significant differences in knowledge, behavior, and practices impacted by IPAC training throughout the geographical span of respondents.

There was a distinctive almost even pattern of the current professional level among our respondents. They belonged to the two extreme levels, 43% were in their early career, followed by 35% either being professors or consultants, and the least 24% were at their intermediate career. In previously published reports the respondents were mostly between 1-5 years’ experience category 66%^11^, 72%^18^, and 64%^23^. There were no significant differences observed between the professional levels of our physicians.

When asked about ever being infected with COVID-19, more than 14% of the respondents said they were tested positive for the disease. Up to mid-June 2020 in Ontario, Canada 5815 HCW were confirmed to have COVID-19^24^. In Bangladesh, till August 10, 2020, a total of 7442 health workers have been infected with the coronavirus, among them, 2542 were physicians with 105 deaths^25^.

The majority of respondents in the present study sought medical/scientific information about COVID-19 from national/international guidelines like-WHO/CDC/local ministry of health (>86%). Similar findings were reported before^26^, however, many articles have denoted that majority of their respondents relied on unverified resources like social media, web blogs, word of mouth, and so on^18, 19, 23, 27^. Since most of our respondents obtained information from international or national official sources, this signifies the validity of their information, which ensures proper behavior and practice pertaining to infection prevention and control.

Receiving IPAC training for COVID-19 was very high (72%) among our respondents. The majority (60%) of the training were from their workplace, and 21% from WHO/CDC. About 50% of the responding physicians received their training both online and in-person and 62% of those training sessions were for more than 3 hours. Despite reasonable searches, we did not find any studies revealing the information about IPAC training and its correlation with knowledge, behavior, and practice of physicians or HCWs.

In the knowledge domain, most of our respondents could correctly choose the right answers. The majority of our participants showed high knowledge scores and they were considered either as competent (40%) or as advanced (43%). Receiving training influenced the knowledge score considerably, which also varied significantly among the knowledge categories. About 70% of the respondent scored more than 6 out of 8 and 9% scored 100% correct. Considering the cut-off score of 4, more than 96% of the respondents had good knowledge of COVID-19. The reason behind this high knowledge score could be because the study was conducted towards the later phase of the COVID-19 pandemic, as well as receiving IPAC training from different sources.

Knowledge is an important factor that directly affects behavior and practice^11, 19^. High levels of knowledge score in our study are in keeping with high confidence levels among the physicians. Most of our respondents showed positive behavior (69%) and good practices (60%) towards COVID-19. This trend was higher in the cohort that received IPAC training. The relationship between higher knowledge and better behavior/practices were reported before by other studies^18, 26-28^.

### Strength

To our knowledge, this is the earliest report of training effectiveness on knowledge, behavior, and practice of physicians regarding COVID-19. The questionnaire was designed based on the information available on WHO, CDC, and national regulatory websites. Moreover, a dual validation was performed to increase the reliability of the study results.

### Limitations

As convenience sampling was used, the results could not be generalizable. Since it was an asynchronous, web-based, cross-sectional survey, there are fair chances for the participants to seek the correct answers online before responding.

### Future Directions

Considering the training effectiveness pattern of our study, a robust, multicentre, and systematic experiment could be designed to better understand the significance of training modules.

## 5. Conclusion

The unprecedented global pandemic calls for substantial awareness about the novel coronavirus. Particularly, physicians as the leader of combating this challenge should have extensive readiness to prevent and manage COVID-19 efficiently. Proper information and data need to be cascaded from the authentic sources to avoid the ongoing jeopardy around this novel contagion. Our study finding highlights that IPAC training is effective in enhancing the knowledge of the doctors. We also revealed improvement in behavior and practices in response to the training.

Therefore, we strongly recommend to design and implement appropriate training modules for the physicians as well as other HCW to alleviate the waves of COVID-19 and other outbreaks to come.

## Data Availability

Yes

## Notes

### Competing Interest Statement

The authors have declared no competing interest.

### Funding Statement

no external funding was received

### Author Declarations

Ethical approval was obtained from Holy Family Red Crescent Medical College, Dhaka, Bangladesh. Ref: HFRCMC-IRC/2020.09.050

